# Persistence of endogenous SARS-CoV-2 and pepper mild mottle virus RNA in wastewater settled solids

**DOI:** 10.1101/2022.01.06.22268855

**Authors:** Laura Roldan-Hernandez, Katherine E. Graham, Dorothea Duong, Alexandria B. Boehm

**Affiliations:** Department of Civil and Environmental Engineering, Stanford University, 473 Via Ortega, Stanford 94305, California, United States; Verily Life Sciences, South San Francisco, 94080, California, United States

**Keywords:** SARS-CoV-2, PMMoV, decay, settled solids, wastewater

## Abstract

Limited information is available on the decay rate of endogenous SARS-CoV-2 and pepper mild mottle virus (PMMoV) RNA in wastewater and primary settled solids, potentially limiting an understanding of how transit or holding times within wastewater infrastructure might impact RNA measurements and their relationship to community COVID-19 infections. In this study, primary settled solids samples were collected from two wastewater treatment plants in the San Francisco Bay Area. Samples were thoroughly mixed, aliquoted into subsamples, and stored at 4°C, 22°C, and 37 °C for 10 days. The concentration of SARS-CoV-2 (N1 and N2 targets) and PMMoV RNA was measured using an RT-ddPCR. Limited decay (< 1 log_10_ reduction) was observed in the detection of viral RNA targets at all temperature conditions, suggesting that SARS-CoV-2 and PMMoV RNA can be highly persistent in solids. First-order decay rate constants ranged from 0.011 - 0.098 day^-1^ for SARS-CoV-2 RNA and 0.010 - 0.091 day^-1^ for PMMoV RNA, depending on temperature conditions. Slower decay was observed for SARS-CoV-2 RNA in primary settled solids compared to previously reported decay in wastewater influent. Further research is needed to understand if solid content and wastewater characteristics might influence the persistence of viral RNA targets.

**Synopsis:** SARS-CoV-2 and PMMoV genomic RNA is highly stable in wastewater settled solids over 10 days at several environmentally relevant temperatures.

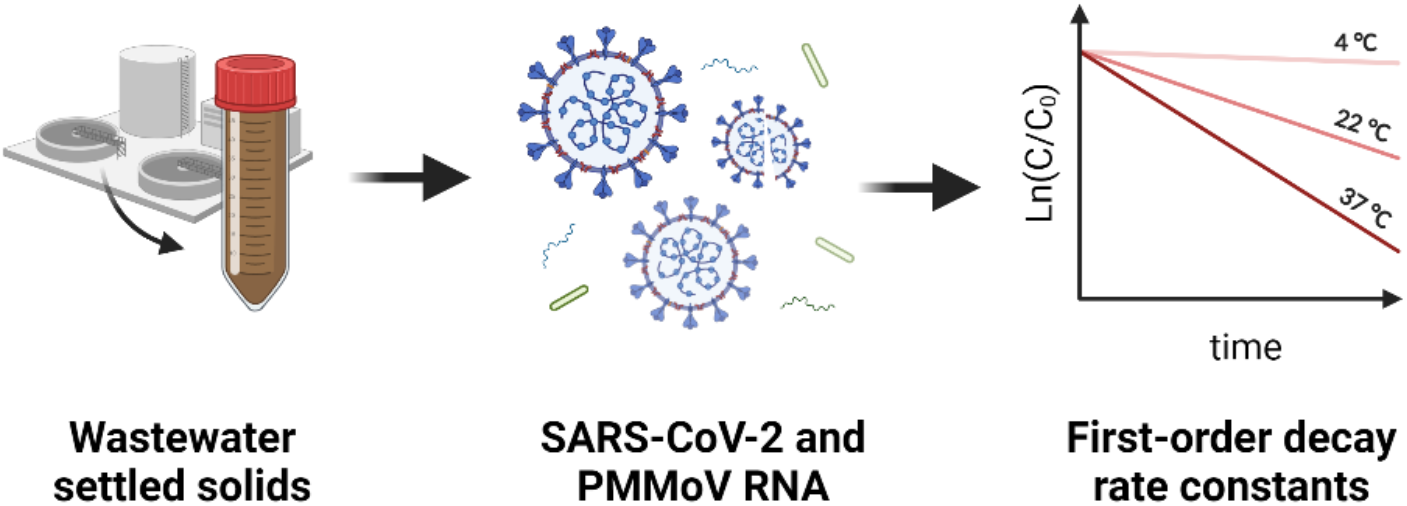

## Introduction

Previous studies showed that RNA from SARS-CoV-2 is naturally concentrated in primary settled solids at wastewater treatment plants.^1,2^ Further, researchers have found a significant correlation between SARS-CoV-2 RNA isolated from primary settled solids samples and laboratory-confirmed COVID-19 cases ^3–7^ thus, settled solids might be an advantageous medium for wastewater-based epidemiology efforts. Laboratory-confirmed COVID-19 incidence may be considerably smaller than actual COVID-19 incidence owing to test-seeking behaviors which are influenced by testing availability and symptom presence and severity.^8,9^ Some studies aim to infer actual COVID-19 incident cases from wastewater concentrations of SARS-CoV-2 RNA using models.^10–12^ Such models require the input of various parameters including fecal loads, viral shedding rates, flow to the wastewater treatment plant, decay rate constant and partition coefficient of SARS-CoV-2 RNA, and the average residence time of wastewater prior to sample collection. In some models, PMMoV RNA is used as an endogenous viral process control and as a fecal strength control.^5^ The accuracy of these models is still uncertain and further research is still needed to improve them. Understanding the decay rate of SARS-CoV-2 and PMMoV RNA is critical in their implementation.

Only a few studies ^13–17^ have documented the persistence of SARS-CoV-2 RNA and infectious SARS-CoV-2 viruses in wastewater (Table 1), all those studies have been conducted using wastewater influent as the experimental matrix. Additionally, most of these studies do not measure the decay of endogenous SARS-CoV-2 RNA, rather they seed the wastewater with an exogenous virus. A recent study by Chik et al.^18^ found that surrogate spikes of SARS-CoV-2 (in this case gamma-irradiated inactivated SARS-CoV-2) may exhibit different solid-liquid partitioning behaviors compared to SARS-CoV-2 naturally found in wastewater. Virus persistence can be influenced by the degree of sorption to solids.^19^ To date, no study has documented the decay of SARS-CoV-2 RNA in wastewater primary settled solids, based on a systematic search of the literature on 11 November 2021 (Table S1).

**Table 1:** First-order decay rate constants (k) and time needed to achieve 90% reduction (T_90_) of SARS-CoV-2 and PMMoV in wastewater and environmental waters stored at 4-37°C. Papers included were identified in the systematic review described in the SI.

| Reference | Target | Matrix | Sample preparation | Temp. (°C) | k (days<sup>-1</sup>) | T<sub>90</sub> (days) |
| --- | --- | --- | --- | --- | --- | --- |
| Ahmed et al. 2020 | SARS-CoV-2 RNA (N1 gene) | wastewater influent | Samples were spiked with gamma-irradiated SARS-CoV-2 (hCoV-19/Australia/VIC01/2020) | 4<br>15<br>25<br>37 | 0.084<br>0.114<br>0.183<br>0.286 | 27.8<br>20.4<br>12.6<br>8.04 |
|  | SARS-CoV-2 RNA (N1 gene) | wastewater influent | Samples were autoclaved and spiked with gamma-irradiated SARS-CoV-2 (hCoV-19/Australia/VIC01/2020) | 4<br>15<br>25<br>37 | 0.054<br>0.077<br>0.171<br>0.405 | 42.6<br>29.9<br>13.5<br>5.7 |
| Bivins et al. 2020 | SARS-CoV-2 RNA (E gene) | wastewater influent | Samples were immediately frozen after collection, then thawed and spiked with SARS-CoV-2 (nCoV-WA1-2020; MN985325.1) | 20 | 0.09, 0.67 | 25.6, 3.4 |
|  | Infectious SARS-CoV-2 virus | wastewater influent | Samples were immediately frozen after collection, then thawed and spiked with SARS-CoV-2 (nCoV-WA1-2020; MN985325.1) | 20 | 1.1, 1.4 | 2.1, 1.6 |
| Hokajärvi et al. 2021 | SARS-CoV-2 RNA (E gene) | wastewater influent | Samples were spiked with SARS-CoV-2 (strain not specified). | 4 | 0.04 | 52 |
|  | SARS-CoV-2 RNA (N2 gene) | wastewater influent | Samples were spiked with SARS-CoV-2 (strain not specified). | 4 | 0.06 | 36 |
| Weidhaas et al. 2021 | SARS-CoV-2 RNA (N1 and N2 gene) | wastewater influent | Study conducted with endogenous SARS-CoV-2. | 4<br>10<br>35 | 0.96, 2.16<br>2.16<br>4.32 | 2.4, 1.1<br>1.1<br>0.5 |
| Oliveira et al. 2021 | Infectious SARS-CoV-2 virus | wastewater influent | Samples were autoclaved and spiked with SARS-CoV-2 (SARS.CoV2/SP02.2020.HIAE.Br) | 4<br>24 | 0.19<br>0.83 | 12.1<br>2.9 |
|  | Infectious SARS-CoV-2 virus | wastewater influent | Samples were autoclaved, filtered through a 0.22 µm membrane, and spiked with SARS-CoV-2 (SARS.CoV2/SP02.2020.HIAE.Br) | 24 | 0.80 | 2.8 |
| Rachmadi et al. 2016 | PMMoV RNA | wetland water | Study conducted with endogenous PMMoV. | 4<br>25<br>37 | 0.04<br>0.05<br>0.08 | 57.6<br>46.1<br>28.8 |

Normalizing SARS-CoV-2 RNA concentrations by concentrations of PMMoV is a widespread practice for relating SARS-CoV-2 RNA target concentrations to laboratory-confirmed COVID-19 incidence. Normalizing by PMMoV theoretically controls for fluctuations in population contributing to a given wastewater sample; however, it may also correct for variation in RNA recoveries between samples^5^, and for losses of RNA during storage.^20^ PMMoV is a non-enveloped single-strand RNA virus highly abundant in human feces and wastewater.^21,22^ Studies have shown that PMMoV RNA is remarkably stable in wastewater and exhibits almost no seasonal variation.^23^ Rachmadi et al.^24^ studied the persistence of endogenous PMMoV RNA using RT-qPCR in constructed wetlands and found limited decay at three temperatures (4 °C, 22 °C, and 37 °C) for 21 days, suggesting it is highly persistent. Given that PMMoV RNA concentrations are used in wastewater monitoring programs to normalize SARS-CoV-2 RNA concentrations, studying its persistence relative to SARS-CoV-2 targets is important. To date, no study has measured the persistence of PMMoV RNA in wastewater or primary settled solids.

In this study, we measured the first-order decay rate constant of SARS-CoV-2 (N1 and N2 targets) and PMMoV RNA in primary settled solids from two different wastewater treatment plants. Primary settled solids samples were stored at three temperatures (4°C, 22°C, and 37°C), which represent typical environmental conditions from cold, temperate, and tropical regions. We also assessed the effects of temperature and wastewater treatment plant on the decay rate constants and compared the decay rate constants for SARS-CoV-2 RNA in primary settled solids to previously reported decay rate constants in wastewater.

### Materials & Methods

#### Study area and sample collection

Primary settled solids samples were collected from the San José-Santa Clara Regional Wastewater Facility (POTW A) and Sacramento Regional Wastewater Treatment Plant (POTW B) on August 9, 2021. POTW A is an advanced-secondary treatment plant located in Santa Clara County, California, USA. The plant serves approximately 1.4 million people and processes an average flow of 110 million gallons per day (MGD). The residence time in the sewer network is around 4-18 hours. Approximately 10 mg/L of FeCl_3_ is added prior to the headworks for odor control. The residence time of solids in the primary clarifier is estimated to be 1-2 hours. POTW B is a secondary treatment plant located in Sacramento County, California, USA. The plant serves approximately 1.6 million people and processes an average flow of 124 MGD. The residence time in the sewer network is approximately 15 hours and the residence time of solids in the primary clarifier is estimated to be 1 hour.

A 24-hour composite sample (500 mL gathered every 4 hours) was collected from the primary sludge line of POTW A. For POTW B, a 2 L grab sample was collected from the primary sludge line. Both samples were collected in 10% HCl acid-washed plastic containers and stored on ice during transportation to the laboratory.

#### Sample processing

Sample processing began within 24 hours of sample collection. Samples from each POTW were thoroughly mixed and aliquoted into 50 mL conical tubes, for a total of 32 subsamples per POTW. Each time point had a biological replicate. Subsamples were then placed inside opaque boxes (to shield samples from light) and placed into constant temperature rooms at 4°C, 22°C, and 37°C. Two subsamples were processed immediately to determine the initial concentrations of PMMoV and SARS-CoV-2 RNA at time zero (t=0). Another subsample was reserved for percent solids analysis which required heating the sample at 105°C for 24 hours and comparing the weight before and after drying. Table S3 includes the percent of solids measured in primary settled solid samples collected from each POTW.

On days 2, 4, 6, 8, and 10, subsamples were retrieved from each temperature condition. Each 50 mL conical tube was centrifuged at 4,200 rpm for 40 minutes at 4°C. The supernatant was decanted and about 0.225 g of solids were aliquoted into 15 ml conical tubes. Solids were immediately resuspended with 3 mL of DNA/RNA Shield (Zymo Research; cat. no. R1100-250) to prevent further degradation and preserve the genetic integrity of the subsamples. The final concentration of dewatered solids resuspended in spiked DNA/RNA Shield was approximately 75 mg/ml. This concentration of solids was chosen as it was found to alleviate inhibition in the downstream RT-ddPCR.^25^ All subsamples were spiked with 4.5 μL of bovine coronavirus vaccine (Zoetis; #CALF-GUARD) to estimate viral RNA extraction efficiencies. RNA extracts needed to have at least a 10% recovery efficiency to be included in the analysis, otherwise they were excluded from the data analysis.

#### RNA extraction

RNA was extracted from subsamples in two batches. Subsamples in Batch 1 (t = 0, 2, and 4) were processed on 13 August 2021 and Batch 2 (t = 6, 8, and 10) on 20 August 2021. RNA extraction was performed following the high throughput RNA extraction and PCR inhibitor removal protocol for settled solids.^26^ Briefly, RNA was extracted from 300 μl of a homogenized sample using the Chemagic™ Viral DNA/RNA 300 Kit H96 for the Perkin Elmer Chemagic 360 followed by PCR Inhibitor Removal with the Zymo OneStep-96 PCR Inhibitor Removal Kit. Two extraction replicates were completed for each subsample. Extraction negative controls consisted of DNase/RNase-free water. Extraction positive controls consisted of DNA/RNA Shield solution spiked with SARS-CoV-2 genomic RNA (ATCC® VR-1986D™) and bovine coronavirus (BCoV). Each RNA extract was aliquoted into separate 1.5 ml DNA LoBind tubes and stored at - 80°C until quantification; samples were stored for approximately 10 days.

#### ddRT-PCR

The concentration of SARS-CoV-2 N1 and N2, PMMoV, and BCoV was determined in each subsample. N1 and N2 were quantified in a duplex assay using the One-Step RT-ddPCR Advanced kit for probes (BioRad; cat. no. 1864021). Each 22-μL reaction consisted of 1X BioRad Supermix, 1X reverse transcriptase, 15 mM DTT, 0.9 μM of each primer, 0.25 μM of each probe, and 5.5 μL template RNA. Primers and probes are listed in Table S2. An individual RNA extract was used for the N1/N2 assay. RNA extracts were run undiluted and six technical replicates (6 ddRT-PCR wells) were merged for analysis. Negative (Mastermix + DNase/RNase-free water) and positive (SARS-CoV-2 genomic RNA ATCC® VR-1986D™) RT-ddPCR controls were included on each plate. Negative and positive extraction controls were also processed for each RNA extraction batch.

A separate RNA extract (extraction replicate) was used for the PMMoV/BCoV assays to prevent freeze-thaw effects in the detection of RNA targets. The duplex assay was performed using the One-Step RT-ddPCR Advanced kit for probes (BioRad; cat. no. 1864021) consisting of 1X BioRad Supermix, 1X reverse transcriptase, 15 mM of dithiothreitol (DTT), 0.9 μM of each primer, 0.4 μM of PMMoV probe, 0.25 μM of BCoV probe, and 5.5 μL template RNA. RNA extracts were diluted 1:100 before ddRT-PCR and two technical replicates (2 ddRT-PCR wells) were merged for analysis. Negative controls (Mastermix + DNase/RNase-free water) and positive controls (a mixture of PMMoV oligos (IDT) and BCoV amplicons generated from end-point PCR) were included on each RT-ddPCR plate. Negative and positive extraction controls were also processed for each RNA extraction batch. Results from the extraction positive control (DNA/RNA Shield spiked with BCoV) were used to estimate the extraction efficiency for each subsample.

Reagents were pipetted into 96-well plates, sealed using a PX1 PCR plate sealer, and vortexed for 30 seconds. Droplets were generated using an AutoDG (BioRad) using automated droplet generation oil for probes (BioRad; cat. no.1864110). Once droplets were generated, plates were sealed and placed onto a thermocycler within 30 minutes of generation. N1/N2 assay plates were thermocycled as follows: 50°C for 60 minutes, 95°C for 10 minutes, 40 cycles of 94°C for 30 seconds then 55°C for 1 minute, followed by 98°C for 10 minutes, and 4°C for at least 30 minutes. PMMoV/BCoV plates were thermocycled as follows: 50°C for 60 minutes, 95°C for 10 minutes, 40 cycles of 94°C for 30 seconds then 56°C for 1 minute, followed by 98°C for 10 minutes, and 4°C for at least 30 minutes. Plates were moved to a QX200 droplet reader (BioRad) within 48 hours of thermocycling. RT-ddPCR data was manually thresholded and exported using the QuantaSoft and QuantaSoft Analysis Pro (BioRad) Software. Droplets were visually inspected and classified following the SARS-CoV-2 and BCoV/PMMoV post-processing analysis procedure described in Wolfe et al.^27^ RT-ddPCR outputs were also converted to copies per g of dry weight using dimensional analysis. ^27^

### Data analysis

Dimensional and statistical analyses were performed using Microsoft Excel and Rstudio (version 4.1.2). First-order decay constants (k) were calculated for experiments at each temperature and POTW using the average concentration of SARS-CoV-2 and PMMoV RNA at each time point (day 0, 2, 4, 6, 8, and 10). k and the time needed to achieve a 90% reduction in concentration (T_90_) were calculated using the following equations:

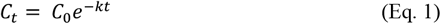

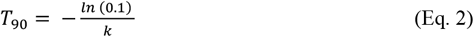

where C_t_ is the concentration of the viral RNA at time t, C_0_ is the concentration of viral RNA at time zero, and k is the first-order decay constant. k was calculated as the slope of ln (C_t_/C_o_) over time using a linear least-squares regression model in R. Goodness of fit of the linear regression model was assessed by determining the coefficient of determination (r^2^) and by visually inspecting the normality and homogeneity of variance of the residuals vs fitted values and Q-Q plots. A Shapiro-Wilk test was also performed to assess the normality assumption.

A multiple linear regression model (Eq. 3) was used to model the means of k as a function of temperature, RNA target, and POTW and to determine which factors had a significant main effect on k. Interaction effects were also included (discussed in results). All analyses were performed in Rstudio using the “lm” function. The initial regression model was then modified to include only factors that had a significant effect on k:

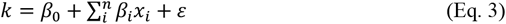

where β_0_ is the intercept, β_i_ is the regression coefficient for each factor x_i_ (temperature, RNA target, and POTW), and ε is the residual standard error of the model. log_10_k values of SARS-CoV-2 RNA from this study were also compared to previously reported log_10_k values of SARS-CoV-2 RNA in wastewater using a linear regression model in R. log_10_k values were used in that analysis because previous work has shown an exponential relationship between k and temperature for viruses in environmental waters. ^28,29^ P<0.05 was used to assess statistical significance.

## Results

### QA/QC

This study takes into consideration the control and process checklist provided in the Environmental Microbiology Minimum Information (EMMI) guidelines^30^. Extraction and RT-ddPCR positive and negative controls were positive and negative, respectively, for all viral RNA targets. The median BCoV recovery was 60% across all primary settled solids samples (POTW A: median = 55% and POTW B: median = 66%). All extractions were above 10% recovery and included in the analysis. BCoV is used solely as a process and gross inhibition control; no attempt was made to correct concentrations by recovery owing to the complexities associated with estimating viral RNA recovery using surrogate viruses.^31^

### Decay rate model

Limited decay (< 1 log_10_ reduction over 10 days) was observed for all RNA targets and at all temperature conditions. Table 2 shows the first-order decay rate constants and time needed to achieve a 90% reduction in concentration for SARS-CoV-2 (N1 and N2 targets) and PMMoV RNA. k was significantly different from 0 for all conditions (p<0.05) and varied from 0.010 to 0.091 day^-1^ depending on the RNA target and temperature conditions. The linear model was a good fit for most decay curves (median r^2^ = 0.60, median RMSE = 0.21; Table 2); however, some experiments had r^2^ less than 0.3. These same experiments had small RMSE and very small k values indicative of limited decay with time. Residual values from the model exhibited a random pattern which suggests that the linear relationship assumption is reasonable.

**Table 2:**
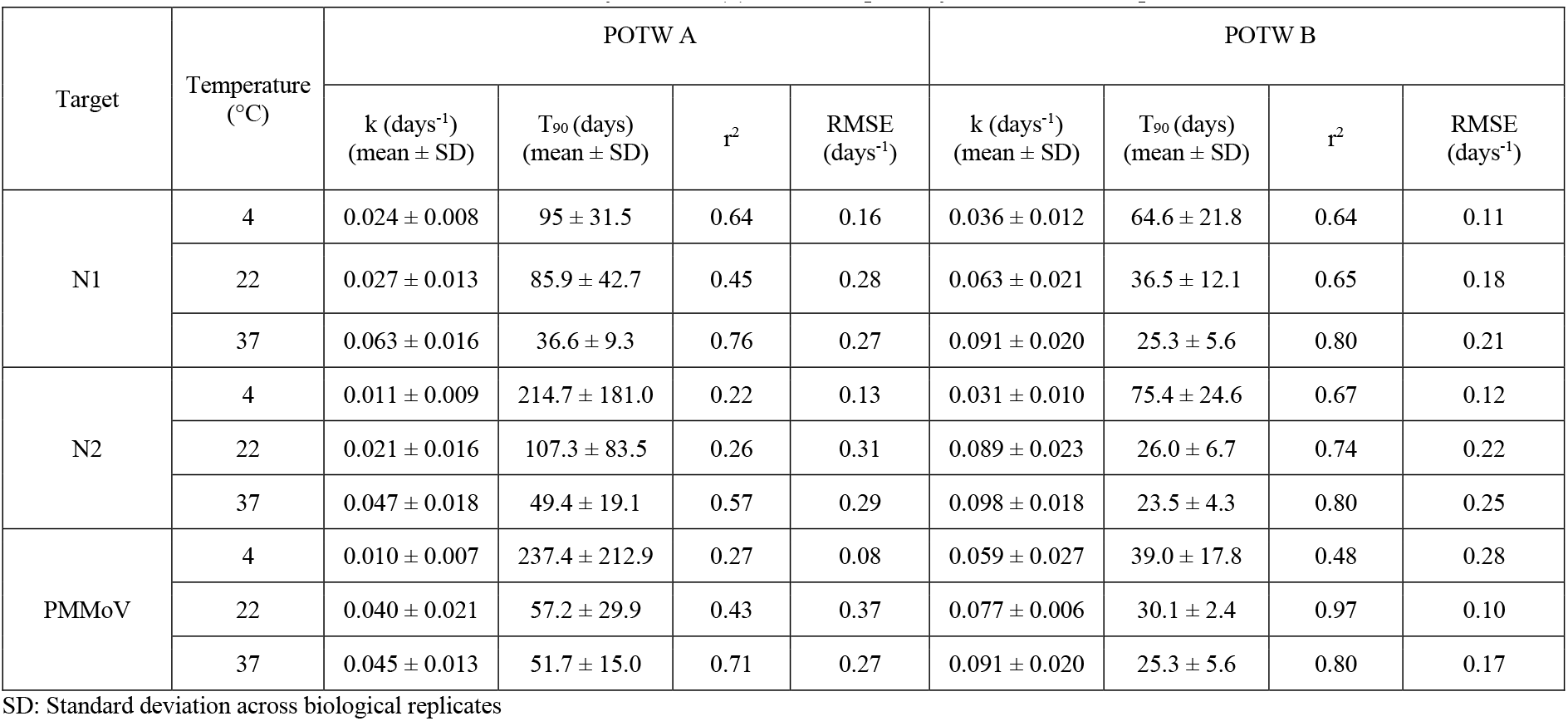
SARS-CoV-2 and PMMoV RNA first-order decay constant (k) and T_90_ in primary settled solids sample from POTW A and POTW B

| Target | Temperature (°C) | POTW A |  |  |  | POTW B |  |  |  |
| --- | --- | --- | --- | --- | --- | --- | --- | --- | --- |
|  |  | k (days<sup>-1</sup>)<br>(mean ± SD) | T<sub>90</sub> (days)<br>(mean ± SD) | r<sup>2</sup> | RMSE<br>(days<sup>-1</sup>) | k (days<sup>-1</sup>)<br>(mean ± SD) | T<sub>90</sub> (days)<br>(mean ± SD) | r<sup>2</sup> | RMSE<br>(days<sup>-1</sup>) |
| N1 | 4 | 0.024 ± 0.008 | 95 ± 31.5 | 0.64 | 0.16 | 0.036 ± 0.012 | 64.6 ± 21.8 | 0.64 | 0.11 |
|  | 22 | 0.027 ± 0.013 | 85.9 ± 42.7 | 0.45 | 0.28 | 0.063 ± 0.021 | 36.5 ± 12.1 | 0.65 | 0.18 |
|  | 37 | 0.063 ± 0.016 | 36.6 ± 9.3 | 0.76 | 0.27 | 0.091 ± 0.020 | 25.3 ± 5.6 | 0.80 | 0.21 |
| N2 | 4 | 0.011 ± 0.009 | 214.7 ± 181.0 | 0.22 | 0.13 | 0.031 ± 0.010 | 75.4 ± 24.6 | 0.67 | 0.12 |
|  | 22 | 0.021 ± 0.016 | 107.3 ± 83.5 | 0.26 | 0.31 | 0.089 ± 0.023 | 26.0 ± 6.7 | 0.74 | 0.22 |
|  | 37 | 0.047 ± 0.018 | 49.4 ± 19.1 | 0.57 | 0.29 | 0.098 ± 0.018 | 23.5 ± 4.3 | 0.80 | 0.25 |
| PMMoV | 4 | 0.010 ± 0.007 | 237.4 ± 212.9 | 0.27 | 0.08 | 0.059 ± 0.027 | 39.0 ± 17.8 | 0.48 | 0.28 |
|  | 22 | 0.040 ± 0.021 | 57.2 ± 29.9 | 0.43 | 0.37 | 0.077 ± 0.006 | 30.1 ± 2.4 | 0.97 | 0.10 |
|  | 37 | 0.045 ± 0.013 | 51.7 ± 15.0 | 0.71 | 0.27 | 0.091 ± 0.020 | 25.3 ± 5.6 | 0.80 | 0.17 |

SD: Standard deviation across biological replicates

### SARS-CoV-2 RNA first-order decay rate constants

The initial average concentrations (mean ± standard deviation) of N1 were 4.48 ± 3.93 and 4.60 ± 3.69 log_10_ copies/g of dry weight at POTW A and POTW B, respectively. For N2, the average initial concentrations were 4.45 ± 3.89 log_10_ copies/g of dry weight at POTW A and 4.55 ± 3.56 log_10_ copies/g of dry weight at POTW B. Figure 1 shows the decay curves of all RNA targets in primary settled solids and Table 2 summarizes the first-order decay constants and T_90_ for all RNA targets and temperature conditions. For N1, k values at POTW A and B were 0.024 and 0.036 day^-1^ at 4 °C, 0.027 and 0.063 day^-1^ at 22 °C, and 0.063 and 0.91 day^-1^ at 37 °C, respectively. At POTW A, the time needed to achieve a 90% reduction in N1 concentration was approximately 95 days at 4 °C, 86 days at 22 °C, and 37 days at 37 °C. N1 T_90_ values at POTW B were approximately 65, 36, and 25 days, depending on temperature conditions. N2 k values at POTW A and B were 0.011 and 0.031 day^-1^ at 4 °C, 0.021 and 0.089 day^-1^ at 22 °C, and 0.047 and 0.98 day^-1^ at 37 °C, respectively. At POTW A, N2 T_90_ values were 215, 107, and 49 days, depending on temperature. At POTW B, N2 T_90_ values were 65, 36, 25 days depending on temperature.

**Figure 1:**
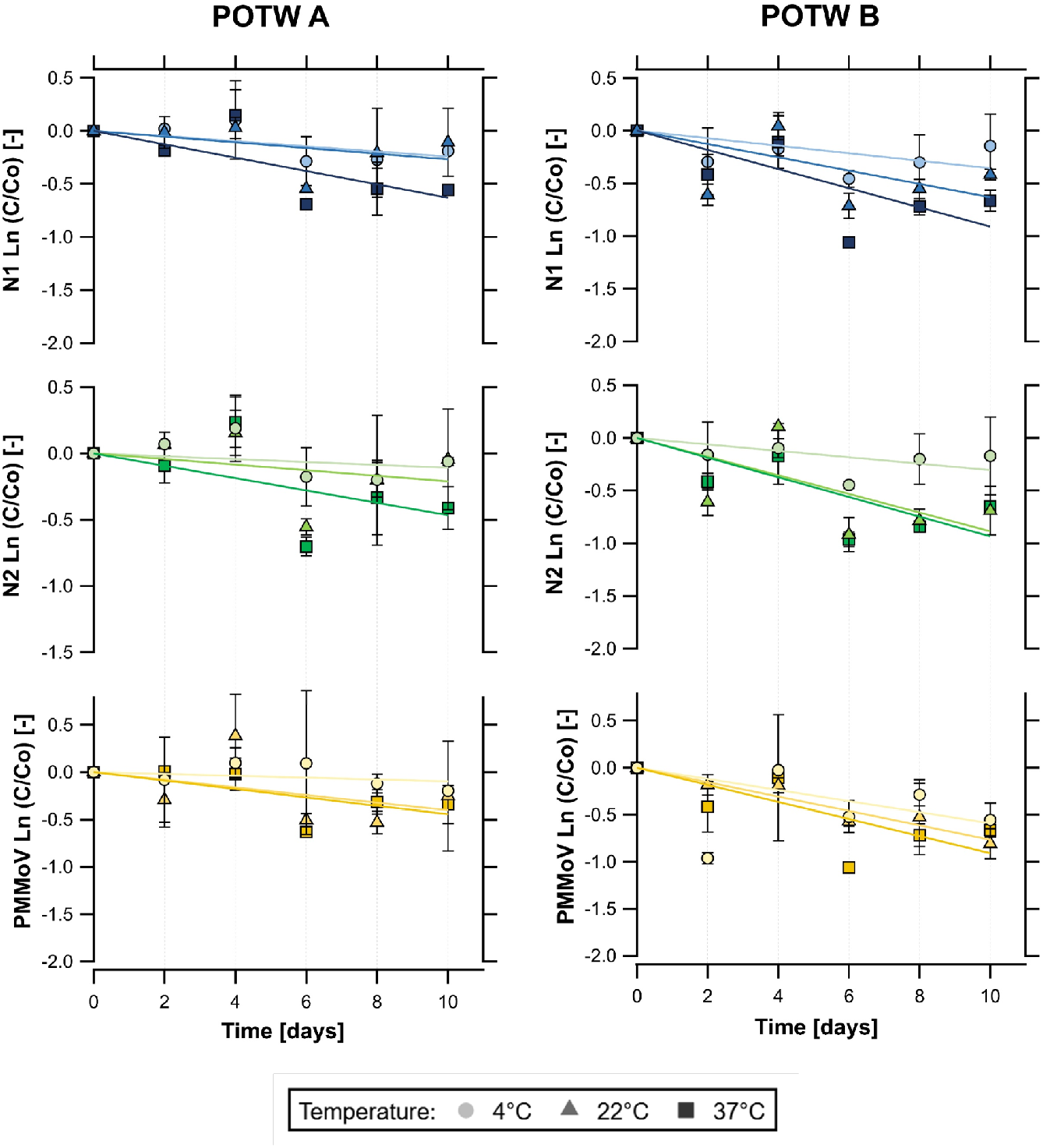
Decay curves of SARS-CoV-2 RNA (N1 and N2) and PMMoV RNA over time (days) in primary settled solids samples stored at 4, 22, and 37 °C. Left column: results from the San José-Santa Clara Regional Wastewater Facility (POTW A). Right column: results from the Sacramento Regional Wastewater Treatment Plant (POTW B). Error bars represent the standard deviation across biological replicates (n = 2).

### PMMoV RNA first-order decay rate constants

The initial mean concentrations (mean ± standard deviation) of PMMoV were 8.32 ± 7.63 and 8.28 ± 7.86 log_10_ copies/g of dry weight at POTW A and POTW B, respectively. Similar to SARS-CoV-2 RNA targets, limited decay was observed for PMMoV RNA at all temperature conditions over 10 days. k values at POTW A and B were 0.010 and 0.059 day^-1^ at 4 °C, 0.040 and 0.077 day^-1^ at 22 °C, and 0.045 and 0.091 day^-1^ at 37 °C, respectively. Based on these results, T_90_ would be 237, 57, and 52 days at POTW A and 39, 30, and 25 days at POTW B depending on temperature conditions.

### Multiple linear regression models for k in primary settled solids

The initial multiple regression model (where k is a function of temperature, RNA target, and POTW) indicates that RNA target (SARS-CoV-2 N1, N2, or PMMoV) is not a significant factor in the model. Temperature and POTW were significant (p<0.05) factors, therefore the regression model was modified to only include these variables, and the interaction term (temperature x POTW) was added to the model. Temperature and POTW remained significant factors in the modified model (*p* =0.0006 and *p* =0.06, respectively) but the interaction term was not significant. The interaction term was therefore removed from the regression model and rerun with just POTW and temperature as factors. The intercept (β_0_) and coefficients (β_temp_ and β_POTW_) of the final regression model were 0.004 per day, 0.001 per day per °C,, and 0.039 per day, respectively. The residual standard error of the regression model was 0.01 per day and adjusted r^2^ was 0.87 (*p* <10^−7^). The positive regression coefficients indicate that k increases with temperature (Figure 2), and that k is higher at POTW B compared to POTW A (as POTW A was coded as the reference POTW in equation 3).

**Figure 2:**
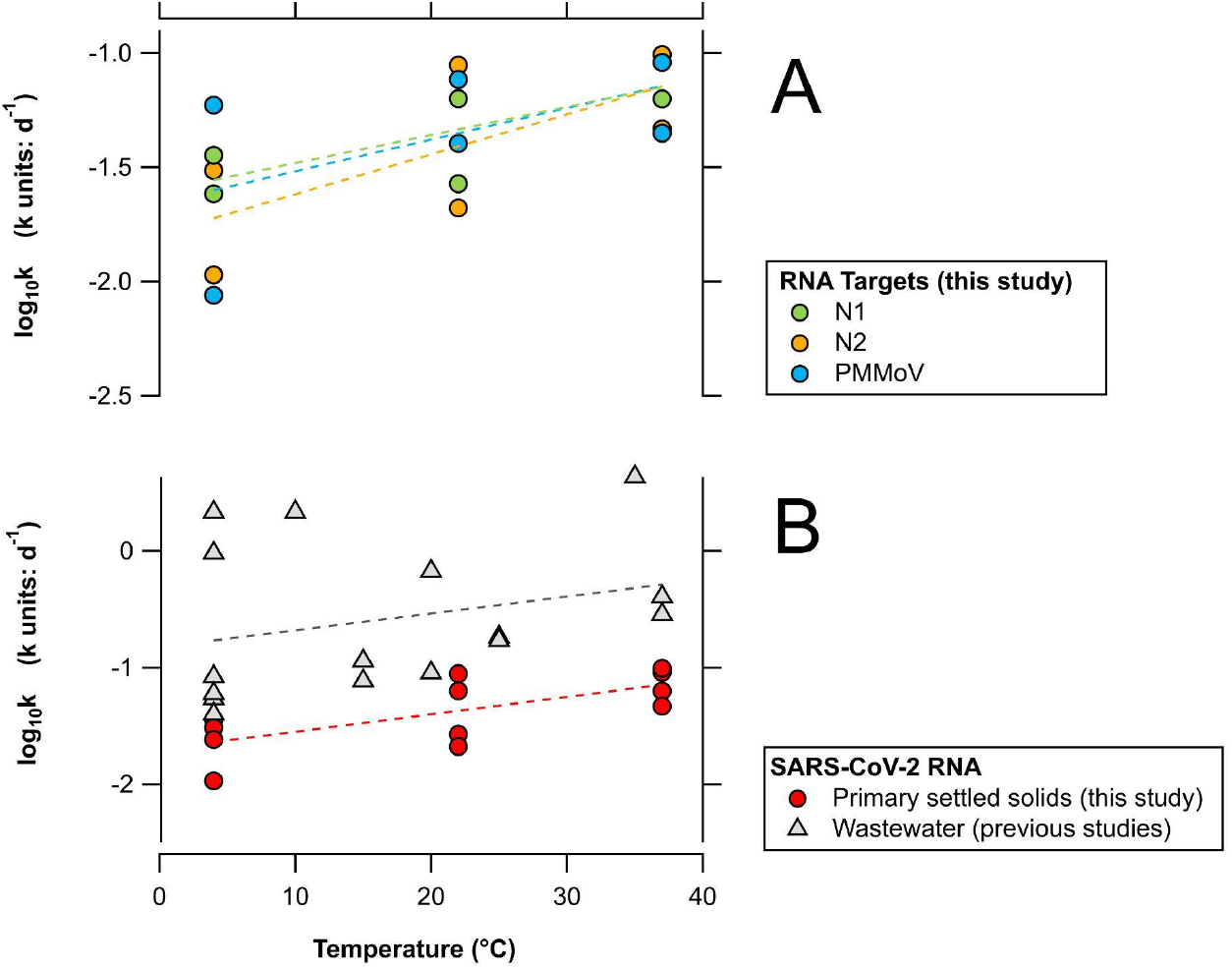
Panel A: log_10_ k of SARS-CoV-2 N1, N2, and PMMoV RNA in primary settled solids from this study stored at 4, 22, and 37 °C. Panel B: log_10_k of SARS-CoV-2 RNA in primary settled solids (this study) and wastewater samples (previous studies) stored at 4-37 °C. Dashed lines represent the linear regression models for each target. Individual k values are summarized in Table 1 (results from previous studies) and Table 2 (results from this study).

### Decay rate of SARS-CoV-2 RNA in primary settled solids versus wastewater

Figure 2 shows log_10_k values of SARS-CoV-2 RNA (N1 and N2 targets) in primary settled solids from this study and previously reported log_10_k values of SARS-CoV-2 RNA (N1, N2, and E targets) in wastewater samples stored at 4 - 37 °C for various durations of time (Table 1). The slope of log_10_k versus temperature was similar for experiments conducted in primary settled solids (this study) and wastewater (0.0149 vs 0.0145). However, a lower y-intercept was observed for primary settled solids compared to wastewater samples (−1.70 vs -0.83). log_10_k values for SARS-CoV-2 RNA in primary settled solids has a positive correlation with temperature (r^2^ = 0.49, *p* = 0.01). However, a poor fit was achieved for the linear regression model that regressed log_10_k values and temperature from various studies that used wastewater influent (r^2^ = 0.07, *p* = 0.29).

## Discussion

SARS-CoV-2 and PMMoV RNA can be highly persistent in primary settled solids. The persistence of viral RNA targets and infectious viruses can be influenced by the physical, chemical, and biological characteristics of wastewater. These factors include temperature, pH, organic matter, dissolved oxygen, solid content, and microbial activity.^19,29,32^ In this study, the decay rate constants of all RNA targets increased with temperature, which is consistent with previous studies of viral decay in the environment.^13–17,24^ However, limited information is available on how solid content in wastewater and primary settled solids can influence the decay rate constants of SARS-CoV-2 and PMMoV RNA. In this study, faster decay rates were observed for RNA targets at POTW B compared to POTW A. Primary settled solid samples from POTW B had a greater percentage of solids (16.57% vs 14.07%) and initial concentration of PMMoV RNA compared to POTW A, which might suggest a higher concentration of fecal matter. Further research is needed to better understand the factors that influence the decay of these targets in the environment.

Decay rate constants of SARS-CoV-2 RNA from this study were compared to previous rate constants of SARS-CoV-2 RNA in wastewater influent. The linear model of log_10_k values of SARS-CoV-2 RNA as a function of temperature displays comparable slopes for experiments conducted using primary settled solids and wastewater influent samples, suggesting that the RNA targets have similar sensitivity to temperature in both matrices. Figure 2 also suggests that SARS-CoV-2 RNA is more persistent in primary settled solids compared to wastewater. In this study, T_90_ values for SARS-CoV-2 RNA ranged from 24 -214 days, depending on temperature conditions and POTW. These values were higher than previously reported T_90_ values for SARS-CoV-2 RNA in wastewater (ranging from 0.5 - 52 days) at similar temperature conditions.

Decay rate constants for N1, N2, and PMMoV RNA targets (all ∼70 bp in length) were not significantly different from each other and quite small. These RNA targets may be located on intact genomes inside of virus capsids and envelopes (for N1 and N2), fragmented genomes inside damaged capsids, or exterior to the virus capsid. Wurtzer et al.^33^ showed that SARS-CoV-2 RNA can exist in diverse forms in wastewater whereas Robinson et al.^34^ indicate that SARS-CoV-2 targets are likely to be present within lipid envelopes in wastewater. Other studies have examined the persistence of viral RNA targets in the environment and generally found the RNA targets are more persistent than infectious viruses as measured using host cell lines.^14,29^ While RNA is generally believed to be quite labile, Walters et al.^35^ showed that the RT-PCR target of naked enterovirus RNA was persistent in seawater microcosms and present for up to 15 days. Research is needed to better understand RNA persistence in the environment, especially as molecular biology methods for quantifying viral nucleic acids in the environment become more accessible and affordable for public health surveillance.

It should be noted that SARS-CoV-2 has not been cultured from raw wastewater^34^ therefore the scientific evidence to date indicates that detection of RNA in these samples is not associated with infectious viral particles. Moreover, the decay rate constants reported herein are not to be misinterpreted as decay rate constants for infectious SARS-CoV-2. Bivins et al.^14^ measured decay rate constants of infectious SARS-CoV-2 seeded into wastewater and found T_90_ = 1.6 to 2.1 days; they observed longer persistence of SARS-CoV-2 RNA in the same experiment T_90_= 3.3 to 26.2 days.

This is the first study that estimates the decay rate of PMMoV RNA in primary settled solids or any wastewater-related matrix. PMMoV is naturally found in human feces, wastewater, and primary settled samples and previous studies have found that the molecular signal of PMMoV is remarkably temporally stable in wastewater with almost no seasonal variation throughout the year.^23^ For example, Rachmadi et al.^24^ studied the persistence of endogenous PMMoV RNA in constructed wetlands and observed limited decay at 4 °C, 22 °C, and 37 °C for over 21 days. Interestingly, the k values reported in constructed wetlands are similar to those reported here for settled solids; 0.04-0.08 days^-1^ in wetland waters and 0.01-0.09 days^-1^ in primary settled solids samples stored at 4-37°C.

At the moment, most SARS-CoV-2 RNA decay experiments in the literature have spiked wastewater samples with different strains of SARS-CoV-2 and in some instances, inactivated the external source of SARS-CoV-2 using gamma-radiation as a safety precaution. This was necessary to ensure a high initial concentration in wastewater samples, so that decay could be followed. Only one previous study^17^ measured the decay rate of endogenous SARS-CoV-2 RNA in wastewater, with T_90_ values ranging from 0.5 – 2.4 days for wastewater influent samples stored at 4-35°C. These results are noticeably lower compared to previous experiments conducted using surrogate spikes of SARS-CoV-2 that reported T_90_ values ranging from 3.4-52 days for samples stored at 4-37°C. Limited information is available on how the decay rate constants of endogenous SARS-CoV-2 and spiked SARS-CoV-2 surrogates differ in wastewater and primary settled solid samples. Chik et al.^18^ found that surrogate spikes of SARS-CoV-2 may exhibit different solid-liquid partitioning behaviors compared to endogenous SARS-CoV-2. Therefore, spiking samples with an external source of SARS-CoV-2 may not necessarily represent natural conditions of SARS-CoV-2 present in wastewater and primary settled solids.

Only a few constraints were encountered during the execution of this experiment. After concentrating and dewatering the subsamples, solids were immediately resuspended in DNA/RNA Shield and stored at 4°C for up to 5 days before extraction. Ideally, samples should be processed as soon as possible. However, the DNA/RNA Shield acts to preserve the integrity of the RNA in the solids and prevent its further degradation; and the results of this study suggest negligible decay of RNA targets at 4°C in the absence of the DNA/RNA Shield. Another possible constraint is that limited decay (< 1 log_10_ reduction) was observed in the detection of all RNA targets through this study. The length of the study was selected based on the average hydraulic residence time of the sewer network (<24 hours) and the average residence time of solids in primary clarifiers (<12 hours) in this study. Future studies could consider experimental durations.

## Conclusion

This study fills critical knowledge gaps regarding the persistence of SARS-CoV-2 and PMMoV RNA in wastewater settled solids. We find that SARS-CoV-2 and PMMoV RNA can be highly persistent in primary settled solids for several weeks and even months depending on temperature conditions. Results from this study and previous studies of SARS-CoV-2 RNA in wastewater suggest limited decay of viral nucleic acids during their transit through the sewer network and within the primary clarifiers, as the time scales of transit in these systems are usually less than 48 hours. This conclusion relies on the assumption that the persistence of the RNA targets in the solids obtained from the primary clarifier is similar to that of the RNA targets on solids suspended in wastewater as it transits through the sewage network. The k values reported herein will be particularly useful in models that link SARS-CoV-2 RNA in settled solids to COVID-19 incidence rates in sewersheds, and aid in the interpretation of SARS-CoV-2 RNA concentrations in settled solids for applications in wastewater-based epidemiology. Future experiments should investigate the decay of other targets used for wastewater-based epidemiology including respiratory syncytial virus (RSV).^36^

## Data Availability

All data produced in the present study are available upon reasonable request to the authors
All data produced in the present work are contained in the manuscript

## Acknowledgments

This study was funded by a gift from the CDC Foundation. We acknowledge the following individuals for assistance with wastewater solids collection: Payal Sarkar (SJ), Noel Enok (SJ), and Amy Wong (SJ); Srividhya Ramamoorthy (Sac), Michael Cook (Sac), Ursula Bigler (Sac), James Noss (Sac), and Lisa C. Thompson (Sac). The graphical abstract was created with BioRender.com.

## Supporting Information

Tables S1, S2, and S3 show the systematic review process, primer design criteria, and characteristics of wastewater treatment plants and primary settled solid samples, respectively.

## References

(1) Kim, S.; Kennedy, L.; Wolfe, M.; Criddle, C.; Duong, D.; Topol, A.; White, B. J.; Kantor, R.; Nelson, K.; Steele, J.; Langlois, K.; Griffith, J.; Zimmer-Faust, A.; McLellan, S.; Schussman, M.; Armmerman, M.; Wigginton, K.; Bakker, K.; Boehm, A. SARS-CoV-2 RNA Is Enriched by Orders of Magnitude in Solid Relative to Liquid Wastewater at Publicly Owned Treatment Works; preprint; Infectious Diseases (except HIV/AIDS), 2021. https://doi.org/10.1101/2021.11.10.21266138.

(2) Li, B.; Di, D. Y. W.; Saingam, P.; Jeon, M. K.; Yan, T. Fine-Scale Temporal Dynamics of SARS-CoV-2 RNA Abundance in Wastewater during A COVID-19 Lockdown. Water Research 2021, 197, 117093. https://doi.org/10.1016/j.watres.2021.117093.

(3) Peccia, J.; Zulli, A.; Brackney, D. E.; Grubaugh, N. D.; Kaplan, E. H.; Casanovas-Massana, A.; Ko, A. I.; Malik, A. A.; Wang, D.; Wang, M.; Warren, J. L.; Weinberger, D. M.; Arnold, W.; Omer, S. B. Measurement of SARS-CoV-2 RNA in Wastewater Tracks Community Infection Dynamics. Nat Biotechnol 2020, 38 (10), 1164–1167. https://doi.org/10.1038/s41587-020-0684-z.

(4) D’Aoust, P. M.; Mercier, É.; Montpetit, D.; Jia, J.-J.; Alexandrov, I.; Neault, N.; Baig, A. T.; Mayne, J.; Zhang, X.; Alain, T.; Langlois, M.-A.; Servos, M. R.; MacKenzie, M.; Figeys, D.; MacKenzie, A. E.; Graber, T. E.; Delatolla, R. Quantitative Analysis of SARS-CoV-2 RNA from Wastewater Solids in Communities with Low COVID-19 Incidence and Prevalence. Water Research 2020, 116560. https://doi.org/10.1016/j.watres.2020.116560.

(5) Wolfe, M. K.; Archana, A.; Catoe, D.; Coffman, M. M.; Dorevich, S.; Graham, K. E.; Kim, S.; Grijalva, L. M.; Roldan-Hernandez, L.; Silverman, A. I.; Sinnott-Armstrong, N.; Vugia, D. J.; Yu, A. T.; Zambrana, W.; Wigginton, K. R.; Boehm, A. B. Scaling of SARS-CoV-2 RNA in Settled Solids from Multiple Wastewater Treatment Plants to Compare Incidence Rates of Laboratory-Confirmed COVID-19 in Their Sewersheds. Environ. Sci. Technol. Lett. 2021, 8 (5), 398–404. https://doi.org/10.1021/acs.estlett.1c00184.

(6) Graham, K. E.; Loeb, S. K.; Wolfe, M. K.; Catoe, D.; Sinnott-Armstrong, N.; Kim, S.; Yamahara, K. M.; Sassoubre, L. M.; Mendoza Grijalva, L. M.; Roldan-Hernandez, L.; Langenfeld, K.; Wigginton, K. R.; Boehm, A. B. SARS-CoV-2 RNA in Wastewater Settled Solids Is Associated with COVID-19 Cases in a Large Urban Sewershed. Environ. Sci. Technol. 2021, 55 (1), 488–498. https://doi.org/10.1021/acs.est.0c06191.

(7) Balboa, S.; Mauricio-Iglesias, M.; Rodriguez, S.; Martínez-Lamas, L.; Vasallo, F. J.; Regueiro, B.; Lema, J. M. The Fate of SARS-COV-2 in WWTPS Points out the Sludge Line as a Suitable Spot for Detection of COVID-19. Science of The Total Environment 2021, 772, 145268. https://doi.org/10.1016/j.scitotenv.2021.145268.

(8) Reese, H.; Iuliano, A. D.; Patel, N. N.; Garg, S.; Kim, L.; Silk, B. J.; Hall, A. J.; Fry, A.; Reed, C. Estimated Incidence of Coronavirus Disease 2019 (COVID-19) Illness and Hospitalization—United States, February–September 2020. Clinical Infectious Diseases 2021, 72 (12), e1010–e1017. https://doi.org/10.1093/cid/ciaa1780.

(9) Iuliano, A. D.; Chang, H. H.; Patel, N. N.; Threlkel, R.; Kniss, K.; Reich, J.; Steele, M.; Hall, A. J.; Fry, A. M.; Reed, C. Estimating Under-Recognized COVID-19 Deaths, United States, March 2020-May 2021 Using an Excess Mortality Modelling Approach. The Lancet Regional Health - Americas 2021, 1, 100019. https://doi.org/10.1016/j.lana.2021.100019.

(10) Ahmed, W.; Angel, N.; Edson, J.; Bibby, K.; Bivins, A.; O’Brien, J. W.; Choi, P. M.; Kitajima, M.; Simpson, S. L.; Li, J.; Tscharke, B.; Verhagen, R.; Smith, W. J. M.; Zaugg, J.; Dierens, L.; Hugenholtz, P.; Thomas, K. V.; Mueller, J. F. First Confirmed Detection of SARS-CoV-2 in Untreated Wastewater in Australia: A Proof of Concept for the Wastewater Surveillance of COVID-19 in the Community. Science of The Total Environment 2020, 728, 138764. https://doi.org/10.1016/j.scitotenv.2020.138764.

(11) Saththasivam, J.; El-Malah, S. S.; Gomez, T. A.; Jabbar, K. A.; Remanan, R.; Krishnankutty, A. K.; Ogunbiyi, O.; Rasool, K.; Ashhab, S.; Rashkeev, S.; Bensaad, M.; Ahmed, A. A.; Mohamoud, Y. A.; Malek, J. A.; Abu Raddad, L. J.; Jeremijenko, A.; Abu Halaweh, H. A.; Lawler, J.; Mahmoud, K. A. COVID-19 (SARS-CoV-2) Outbreak Monitoring Using Wastewater-Based Epidemiology in Qatar. Science of The Total Environment 2021, 774, 145608. https://doi.org/10.1016/j.scitotenv.2021.145608.

(12) Gerrity, D.; Papp, K.; Stoker, M.; Sims, A.; Frehner, W. Early-Pandemic Wastewater Surveillance of SARS-CoV-2 in Southern Nevada: Methodology, Occurrence, and Incidence/Prevalence Considerations. Water Research X 2021, 10, 100086. https://doi.org/10.1016/j.wroa.2020.100086.

(13) Ahmed, W.; Bertsch, P. M.; Bibby, K.; Haramoto, E.; Hewitt, J.; Huygens, F.; Gyawali, P.; Korajkic, A.; Riddell, S.; Sherchan, S. P.; Simpson, S. L.; Sirikanchana, K.; Symonds, E. M.; Verhagen, R.; Vasan, S. S.; Kitajima, M.; Bivins, A. Decay of SARS-CoV-2 and Surrogate Murine Hepatitis Virus RNA in Untreated Wastewater to Inform Application in Wastewater-Based Epidemiology. Environmental Research 2020, 191, 110092. https://doi.org/10.1016/j.envres.2020.110092.

(14) Bivins, A.; Greaves, J.; Fischer, R.; Yinda, K. C.; Ahmed, W.; Kitajima, M.; Munster, V. J.; Bibby, K. Persistence of SARS-CoV-2 in Water and Wastewater. Environ. Sci. Technol. Lett. 2020, 7 (12), 937–942. https://doi.org/10.1021/acs.estlett.0c00730.

(15) de Oliveira, L. C.; Torres-Franco, A. F.; Lopes, B. C.; Santos, B. S. Á. da S.; Costa, E. A.; Costa, M. S.; Reis, M. T. P.; Melo, M. C.; Polizzi, R. B.; Teixeira, M. M.; Mota, C. R. Viability of SARS-CoV-2 in River Water and Wastewater at Different Temperatures and Solids Content. Water Research 2021, 195, 117002. https://doi.org/10.1016/j.watres.2021.117002.

(16) Hokajärvi, A.-M.; Rytkönen, A.; Tiwari, A.; Kauppinen, A.; Oikarinen, S.; Lehto, K.-M.; Kankaanpää, A.; Gunnar, T.; Al-Hello, H.; Blomqvist, S.; Miettinen, I. T.; Savolainen-Kopra, C.; Pitkänen, T. The Detection and Stability of the SARS-CoV-2 RNA Biomarkers in Wastewater Influent in Helsinki, Finland. Science of The Total Environment 2021, 770, 145274. https://doi.org/10.1016/j.scitotenv.2021.145274.

(17) Weidhaas, J.; Aanderud, Z. T.; Roper, D. K.; VanDerslice, J.; Gaddis, E. B.; Ostermiller, J.; Hoffman, K.; Jamal, R.; Heck, P.; Zhang, Y.; Torgersen, K.; Laan, J. V.; LaCross, N. Correlation of SARS-CoV-2 RNA in Wastewater with COVID-19 Disease Burden in Sewersheds. Science of The Total Environment 2021, 775, 145790. https://doi.org/10.1016/j.scitotenv.2021.145790.

(18) Chik, A. H. S.; Glier, M. B.; Servos, M.; Mangat, C. S.; Pang, X.-L.; Qiu, Y.; D’Aoust, P. M.; Burnet, J.-B.; Delatolla, R.; Dorner, S.; Geng, Q.; Giesy, J. P.; McKay, R. M.; Mulvey, M. R.; Prystajecky, N.; Srikanthan, N.; Xie, Y.; Conant, B.; Hrudey, S. E. Comparison of Approaches to Quantify SARS-CoV-2 in Wastewater Using RT-QPCR: Results and Implications from a Collaborative Inter-Laboratory Study in Canada. Journal of Environmental Sciences 2021, 107, 218–229. https://doi.org/10.1016/j.jes.2021.01.029.

(19) Jin, Y.; Flury, M. Fate and Transport of Viruses in Porous Media. In Advances in Agronomy; Elsevier, 2002; Vol. 77, pp 39–102. https://doi.org/10.1016/S0065-2113(02)77013-2.

(20) Simpson, A.; Topol, A.; White, B.; Wolfe, M. K.; Wigginton, K.; Boehm, A. B. Effect of Storage Conditions on SARS-CoV-2 RNA Quantification in Wastewater Solids; preprint; Infectious Diseases (except HIV/AIDS), 2021. https://doi.org/10.1101/2021.05.04.21256611.

(21) Rosario, K.; Symonds, E. M.; Sinigalliano, C.; Stewart, J.; Breitbart, M. Pepper Mild Mottle Virus as an Indicator of Fecal Pollution. Appl Environ Microbiol 2009, 75 (22), 7261–7267. https://doi.org/10.1128/AEM.00410-09.

(22) Symonds, E. M.; Nguyen, K. H.; Harwood, V. J.; Breitbart, M. Pepper Mild Mottle Virus: A Plant Pathogen with a Greater Purpose in (Waste)Water Treatment Development and Public Health Management. Water Research 2018, 144, 1–12. https://doi.org/10.1016/j.watres.2018.06.066.

(23) Kitajima, M.; Sassi, H. P.; Torrey, J. R. Pepper Mild Mottle Virus as a Water Quality Indicator. npj Clean Water 2018, 1 (1), 19. https://doi.org/10.1038/s41545-018-0019-5.

(24) Rachmadi, A. T.; Kitajima, M.; Pepper, I. L.; Gerba, C. P. Enteric and Indicator Virus Removal by Surface Flow Wetlands. Science of The Total Environment 2016, 542, 976–982. https://doi.org/10.1016/j.scitotenv.2015.11.001.

(25) Huisman, J. S.; Scire, J.; Caduff, L.; Fernandez-Cassi, X.; Ganesanandamoorthy, P.; Kull, A.; Scheidegger, A.; Stachler, E.; Boehm, A. B.; Hughes, B.; Knudson, A.; Topol, A.; Wigginton, K. R.; Wolfe, M. K.; Kohn, T.; Ort, C.; Stadler, T.; Julian, T. R. Wastewater-Based Estimation of the Effective Reproductive Number of SARS-CoV-2; preprint; Public and Global Health, 2021. https://doi.org/10.1101/2021.04.29.21255961.

(26) Topol, A.; Wolfe, M.; Wigginton, K.; White, B.; B Boehm, A. High Throughput RNA Extraction and PCR Inhibitor Removal of Settled Solids for Wastewater Surveillance of SARS-CoV-2 RNA V1. https://doi.org/10.17504/protocols.io.btyrnpv6.

(27) Wolfe, M. K.; Topol, A.; Knudson, A.; Simpson, A.; White, B.; Vugia, D. J.; Yu, A. T.; Li, L.; Balliet, M.; Stoddard, P.; Han, G. S.; Wigginton, K. R.; Boehm, A. B. High Frequency, High Throughput Quantification of SARS-CoV-2 RNA in Wastewater Settled Solids at Eight Publicly Owned Treatment Works in Northern California Shows Strong Association with COVID-19 Incidence; preprint; Public and Global Health, 2021. https://doi.org/10.1101/2021.07.16.21260627.

(28) Silverman, A. I.; Boehm, A. B. Systematic Review and Meta-Analysis of the Persistence and Disinfection of Human Coronaviruses and Their Viral Surrogates in Water and Wastewater. Environ. Sci. Technol. Lett. 2020, 7 (8), 544–553. https://doi.org/10.1021/acs.estlett.0c00313.

(29) Silverman, A. I.; Boehm, A. B. Systematic Review and Meta-Analysis of the Persistence of Enveloped Viruses in Environmental Waters and Wastewater in the Absence of Disinfectants. Environ. Sci. Technol. 2021, 55 (21), 14480–14493. https://doi.org/10.1021/acs.est.1c03977.

(30) Borchardt, M. A.; Boehm, A. B.; Salit, M.; Spencer, S. K.; Wigginton, K. R.; Noble, R. T. The Environmental Microbiology Minimum Information (EMMI) Guidelines: QPCR and DPCR Quality and Reporting for Environmental Microbiology. Environ. Sci. Technol. 2021, acs.est.1c01767. https://doi.org/10.1021/acs.est.1c01767.

(31) Kantor, R. S.; Nelson, K. L.; Greenwald, H. D.; Kennedy, L. C. Challenges in Measuring the Recovery of SARS-CoV-2 from Wastewater. Environ. Sci. Technol. 2021, 55 (6), 3514–3519. https://doi.org/10.1021/acs.est.0c08210.

(32) Ho, J.; Stange, C.; Suhrborg, R.; Wurzbacher, C.; Drewes, J. E.; Tiehm, A. SARS-CoV-2 Wastewater Surveillance in Germany: Long-Term PCR Monitoring, Suitability of Primer/Probe Combinations and Biomarker Stability; preprint; Epidemiology, 2021. https://doi.org/10.1101/2021.09.16.21263575.

(33) Wurtzer, S.; Waldman, P.; Ferrier-Rembert, A.; Frenois-Veyrat, G.; Mouchel, J. M.; Boni, M.; Maday, Y.; Marechal, V.; Moulin, L. Several Forms of SARS-CoV-2 RNA Can Be Detected in Wastewaters: Implication for Wastewater-Based Epidemiology and Risk Assessment. Water Research 2021, 198, 117183. https://doi.org/10.1016/j.watres.2021.117183.

(34) Robinson, C. A.; Hsieh, H.-Y.; Hsu, S.-Y.; Wang, Y.; Salcedo, B. T.; Belenchia, A.; Klutts, J.; Zemmer, S.; Reynolds, M.; Semkiw, E.; Foley, T.; Wan, X.; Wieberg, C. G.; Wenzel, J.; Lin, C.-H.; Johnson, M. C. Defining Biological and Biophysical Properties of SARS-CoV-2 Genetic Material in Wastewater. Science of The Total Environment 2022, 807, 150786. https://doi.org/10.1016/j.scitotenv.2021.150786.

(35) Walters, S. P.; Yamahara, K. M.; Boehm, A. B. Persistence of Nucleic Acid Markers of Health-Relevant Organisms in Seawater Microcosms: Implications for Their Use in Assessing Risk in Recreational Waters. Water Research 2009, 43 (19), 4929–4939. https://doi.org/10.1016/j.watres.2009.05.047.

(36) Hughes, B.; Duong, D.; White, B. J.; Wigginton, K. R.; Chan, E. M. G.; Wolfe, M. K.; Boehm, A. B. Respiratory Syncytial Virus (RSV) RNA in Wastewater Settled Solids Reflects RSV Clinical Positivity Rates; preprint; Infectious Diseases (except HIV/AIDS), 2021. https://doi.org/10.1101/2021.12.01.21267014.

