## Supplemental Information for "Persistence of endogenous SARS-CoV-2 and pepper mild mottle virus RNA in wastewater settled solids"

**Number of pages: 5**
**Number of Tables: 3**

**Table S1: Search terms and statistics for literature review; Persistence of SARS-CoV-2 and PMMoV in wastewater or environmental waters.**

| Virus | Search engine and field | Search terms | Number of papers identified | Number of papers included | Number of k extracted | References |
|---|---|---|---|---|---|---|
| SARS-CoV-2 | Web of science core collection<br><br>Search field = title | SARS-CoV-2 AND wastewater AND persistence OR decay OR stability | 9 | 5 | 21 | [1–5] |
| PMMoV | Web of science core collection<br><br>Search field = abstract | PMMoV OR Pepper Mild Mottle Virus AND wastewater or environment* AND persistence OR decay OR stability OR viability | 6 | 1 | 3 | [6] |

k = first-order decay rate constant

**Table S2: Primers and probes for RT-ddPCR assays**

| Assay target | Primer/Probe Sequence (5'-3') | | Aplicon length (bp) | Reference |
|---|---|---|---|---|
| SARS-CoV-2 N1 | Forward | GACCCCAAAATCAGCGAAAT | 72 | Lu et al.[7] |
| | Reverse | TCTGGTTACTGCCAGTTGAATCTG | | |
| | Probe | ACCCCGCATTACGTTTGGTGGACC (5' FAM/ZEN/3' IBFQ) | | |
| SARS-CoV-2 N2 | Forward | TTACAAACATTGGCCGCAAA | 62 | Lu et al. [7] |
| | Reverse | GCG CGACATTCCGAAGAA | | |
| | Probe | ACAATTTGCCCCCAGCGCTTCAG (5' HEX/ZEN/3' IBFQ) | | |
| BCoV | Forward | CTGGAAGTTGGTGGAGTT | 85 | Decaro et al.[8] |
| | Reverse | ATTATCGGCCTAACATACATC | | |
| | Probe | CCTTCATATCTATACACATCAAGTTGTT (5' FAM/ZEN/3' IBFQ) | | |
| PMMoV | Forward | GAGTGGTTTGACCTTAACGTTTGA | 68 | Haramoto et al.[9] and Zhang et al.[10] |
| | Reverse | TTGTCGGTTGCAATGCAAGT | | |
| | Probe | CCTACCGAAGCAAATG (5' HEX/ZEN/3' IBFQ) | | |

**Table S3: Characteristics of POTWs and primary settled solid samples**

| Description | POTW A | POTW B |
|---|---|---|
| Name | San José-Santa Clara Regional Wastewater Facility | Sacramento Regional Wastewater Treatment Plant |
| Population served | 1.4 million | 1.6 million |
| Flow (MGD) | 110 | 124 |
| Average residence time in the sewer network (hours) | 4-18 | 15 |
| Average residence time of solids in the primary clarifier (hours) | 1-2 | 1 |
| Type of sample | 24-hour composite (500 mL gathered every 4 hours) | grab |
| Percent of solid (%) in primary settled solid samples | 14.07 | 16.57 |

(7) Lu, X.; Wang, L.; Sakthivel, S. K.; Whitaker, B.; Murray, J.; Kamili, S.; Lynch, B.; Malapati, L.; Burke, S. A.; Harcourt, J.; Tamin, A.; Thornburg, N. J.; Villanueva, J. M.; Lindstrom, S. US CDC

Real-Time Reverse Transcription PCR Panel for Detection of Severe Acute Respiratory Syndrome Coronavirus 2. *Emerg. Infect. Dis.* **2020**, *26* (8), 1654–1665. https://doi.org/10.3201/eid2608.201246.

(8) Decaro, N.; Elia, G.; Campolo, M.; Desario, C.; Mari, V.; Radogna, A.; Colaianni, M. L.; Cirone, F.; Tempesta, M.; Buonavoglia, C. Detection of Bovine Coronavirus Using a TaqMan-Based Real-Time RT-PCR Assay. *Journal of Virological Methods* **2008**, *151* (2), 167–171. https://doi.org/10.1016/j.jviromet.2008.05.016.

(9) Haramoto, E.; Kitajima, M.; Kishida, N.; Konno, Y.; Katayama, H.; Asami, M.; Akiba, M. Occurrence of Pepper Mild Mottle Virus in Drinking Water Sources in Japan. *Appl. Environ. Microbiol.* **2013**, *79* (23), 7413–7418. https://doi.org/10.1128/AEM.02354-13.

(10) Zhang, T.; Breitbart, M.; Lee, W. H.; Run, J.-Q.; Wei, C. L.; Soh, S. W. L.; Hibberd, M. L.; Liu, E. T.; Rohwer, F.; Ruan, Y. RNA Viral Community in Human Feces: Prevalence of Plant Pathogenic Viruses. *PLoS Biol* **2005**, *4* (1), e3. https://doi.org/10.1371/journal.pbio.0040003.
